# Reactive Postural Responses Predict Risk For Acute Musculoskeletal Injury In Collegiate Athletes

**DOI:** 10.1101/2022.09.09.22279786

**Authors:** Amanda Morris, Nora F. Fino, Ryan Pelo, Daniel M. Cushman, Nicholas E. Monson, Trevor Jameson, Leland E. Dibble, Peter C. Fino

## Abstract

Identifying risk factors for musculoskeletal (MSK) injury is critical to maintain the health and safety of athletes. While current tests consider isolated assessments of function or subjective ratings, objective tests of reactive postural responses, especially when in cognitively demanding scenarios, may better identify risk of MSK injury than traditional tests alone.

**Objectives:** To examine if objective assessments of reactive postural responses, quantified using wearable inertial measurement units, are associated with the risk for acute lower extremity MSK injuries in collegiate athletes.

**Design:** Prospective survival analysis

**Methods:** 191 Division I NCAA athletes completed an instrumented version of a modified Push and Release (I-mP&R) test at the beginning of their competitive season. The I-mP&R was performed with eyes closed under single- and dual-task (concurrent cognitive task) conditions. Inertial measurement units recorded acceleration and angular velocity data that was used to calculate time to stability (TTS). Acute lower extremity MSK injuries were tracked from first team activity for six months. Cox proportional hazard models were used to determine if longer times to stability were associated with faster time to injury.

**Results:** Longer TTS was associated with increased risk of injury; every 250 ms increase in dual-task median TTS was associated with a 37% increased risk of acute, lower-extremity MSK injury.

**Conclusion:** Tests of reactive balance, particularly under dual-task conditions, may be able to identify athletes at risk of acute lower extremity MSK injury. Clinically-feasible, instrumented tests of reactive should be considered in assessments for prediction and prevention of MSK injury in collegiate athletes.

## INTRODUCTION

There is broad interest in consistent, objective screening tools that can predict future acute lower extremity musculoskeletal (MSK) injuries and enable proactive, preventative care. Unfortunately, there are few consistent, objective predictors of future acute lower extremity MSK injury risk that are feasible in a clinical athletic training room setting. Previous research has suggested that the Functional Movement Screen (FMS) ^1^ may be able to identify athletes at risk of future MSK injury. However, the FMS has only a slightly better than 50/50 chance of accurately identifying collegiate athletes at risk of future MSK.^2^ Additionally, common baseline clinical assessments are unable to predict future lower extremity musculoskeletal injury risk.^3^ Drop vertical jumps such as those in, the Landing Error Scoring System (LESS), have been used widely to identify athletes at risk of anterior cruciate ligament injury,^4, 5^ but the ability of drop vertical jumps to predict injury risk has been inconsistent.^5-7^

More sensitive biomechanical assessments of landing and cutting have shown promise in predicting the risk for future acute MSK injuries. For example, knee abduction moment and dynamic valgus measures at landing during a jump-landing task are associated with anterior cruciate ligament injury in female athletes.^6^ But, these assessments may not be feasible in low-budget athletic training rooms, and there remain questions about the association of these measures with prospective injuries.^8^ In addition to biomechanics, cognition appears to play an important role in injury risk. Slower reaction times,^9-11^ slower motor speeds, ^9, 11^ and worse visual and verbal memory scores^9^ from computerized cognitive tests can be associated with future MSK injury, but mixed results^3, 12^ raise doubts about the sole use of cognitive tests to predict acute MSK injury risk.

Recently, more attention has focused on the integration of biomechanics and cognition,^13^ (i.e., neuromuscular control or neuromechanics) as a primary risk factor for acute MSK injury.^14^ Competitive athletics requires processing large amounts of information, integrating sensory stimuli, and executing movements under time constrained conditions.^13, 14^ Thus, the ability to simultaneously execute cognitive and motor tasks, often termed dual-tasks, is critical to safety in sport.^14^ Poor neurocognitive performance has been linked to altered neuromuscular performance such as: increased peak vertical ground-reaction force,^15^ increased knee abduction moment and angle,^15^ and increased peak knee valgus angles.^16^ Alterations in neuromuscular control have been identified as a risk factor for knee^6, 17^ and ankle^17^ MSK injury. Clinical assessments that probe neuromechanical responses, including the identification of a sensory stimuli; integration of multiple senses; and short-, medium-, and long-latency responses to achieve fast, yet precise motion are promising, but under-utilized, avenues for identifying athletes at risk for future acute MSK injury particularly under dual-task conditions.^18, 19^

The purpose of this study was to examine if reactive postural responses (i.e., the neuromechanical responses to a loss of stability) are associated with the risk for acute lower extremity MSK injuries in collegiate athletes.^18^ Reactive postural responses occur when there is a sudden change in posture or support surface, are shaped by the pre-perturbation neuromotor state (i.e. central set), and result from interaction between spinal circuits, the brainstem, and the cerebral cortex.^20^ When exposed to a perturbation, the initial, spinal-mediated response is generally too small to stabilize balance. However, as the response goes on, synergistic muscle activations stabilize balance via mediation from the brainstem and cortex.^20^ Inability of the neuromuscular system to respond quickly or appropriately may result in abnormal joint biomechanics and/or a failure to maintain stability, resulting in a cascading, prolonged response and possible injury.^13^ Previous studies have supported this notion using laboratory-based paradigms; reactive postural responses to the release of a sustained force at the trunk are highly associated with future knee injuries.^21^

Here, we used inertial measurement units (IMUs) to objectively quantify reactive postural responses using an instrumented, modified version of the Push and Release test (I-mP&R), a clinically feasible test that evaluates reactive postural responses.^22^ We hypothesized that collegiate athletes with slower reactive postural responses at a pre-season baseline assessment would be at a higher risk of acute lower extremity MSK injury because of the slower neuromechanical response.^22^

## METHODS

The hypotheses, methods, and procedures of this study were pre-defined in a published protocol (see Phase 2).^18^ Because of protocol deviations due to COVID-19, this current analysis represents a preliminary analysis with minor changes from the protocol. A total of 191 National Collegiate Athletic Association Division I (NCAA D1) collegiate student-athletes, from the University of Utah, across 19 different athletic teams were included (**Table 1**). A comprehensive summary of number of athletes per sport is provided in **Supplementary Table 1**. All participants provided written informed consent under a protocol approved by the University of Utah Institutional Review Board. Inclusion criteria were as follows: (1) 18 to 30 years old and (2) current participant in NCAA D1 athletics. Exclusion criteria were as follows: (1) recent (within 6 months) or planned surgery that would result in future time loss/ practice or competition loss, (2) chronic conditions that could confound testing procedures (i.e. history of vestibular or somatosensory pathology, overuse injuries such as stress fractures), (3) history of major injury to either leg requiring surgery within the last two years, and (4) BMI >40 kg/m^2^. Participants were interviewed about their lifetime concussion and previous musculoskeletal injury history to account for injuries that might have occurred in the previous two years but were before their time as a NCAA athlete and therefore not accounted for in the electronic medical record / injury database. Exclusion based on medical history was determined by the study team.

**Table 1.**
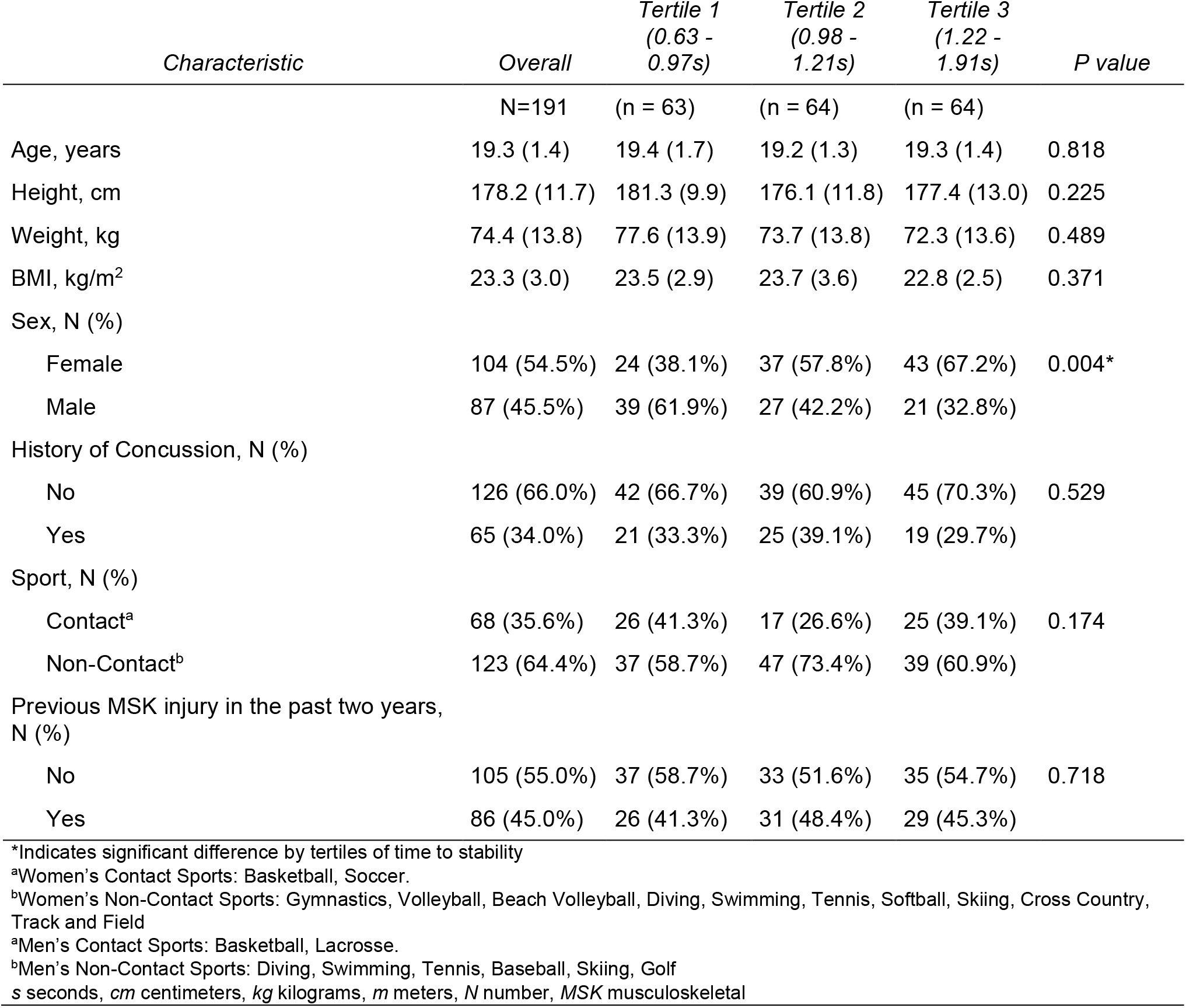
Descriptive Statistics by Tertiles of Dual-Task Time-to-Stability

The instrumented, modified Push and Release test (I-mP&R) was administered as described in Morris et al.^18^ prior to, or at the beginning, of each athlete’s competitive season. IMUs were placed on the participants’ feet, right shank, lumbar spine, and sternum. To determine the time of release, an IMU was placed on the administrator’s hand. At the beginning of each forward and backward trial, a footplate (8′′ long, 5.75′′ wide at the toes, 4′′ wide at the heels) was placed between participants’ feet and removed before beginning the trial to standardize foot placement. Participants were then asked to lean into the administrator’s hands, in a plank-like position with arms at the side, until the administrator judged their center of mass to be just beyond the edge of their base of support. Participants then closed their eyes and the administrator released them unexpectedly. Participants were instructed to do whatever was necessary to regain balance consistent with the prescribed instructions.^18^ The I-mP&R was performed in all four directions (forward, backward, left, and right) and under single and dual-task conditions. Single- and dual-task conditions were presented in a fixed, blocked order. The order of the single- and dual-task blocks was randomized. As soon as participants were in the supported leaning position they were prompted to begin one of the four cognitive tasks in random order: serial subtraction (3’s), phonemic verbal fluency (FAS test), categorical verbal fluency (animal or fruit naming), and reciting every other letter of the alphabet.^23^ Participants responded at least three times before the administrator unexpectedly released the participant. The performance on the cognitive task was not recorded.

Raw linear acceleration and angular velocity data sampled at 128 Hz by IMUs (Opal, APDM Inc, Portland, OR) were used in custom algorithms (MATLAB, MathWorks, Natick, MA) to calculate: time of release, step latency, step length, and time-to-stability (**Supplementary Figure 1**).^22^ The four lean directions were consolidated using the median step length, median time-to-stability, and maximum step latency based on prior recommendations.^22^ The primary measure, as described in the published protocol paper, was median time-to-stability.^18^ Secondary measures included: maximum step latency and median step length.^22^

Acute, lower extremity MSK injuries were tracked from the date of the first organized team activity to March 14, 2020, when athletic activities were halted due to the COVID-19 global pandemic. Injury data was gathered from each participant’s electronic medical records maintained by the University of Utah Athletic Training Staff as part of the Pac-12 Sports Injury Registry Management and Analytics Program / Health Analytics Program (Pac-12 SIRMAP/HAP). Injuries of interest included: any acute orthopedic injury of the lower extremity, pelvis, lumbar spine, or abdomen that resulted in time lost, including but not limited to, joint sprains, musculotendinous strains, and fractures. Any injury resulting in an athlete being unable to fully participate in practice or match play was considered a time-loss injury. Overuse injuries and preexisting conditions were excluded. The date and type of injury was recorded. All statistical analyses were performed with SAS version 9.4 (SAS Institute, Inc., Cary, NC).

Time from first team activity to first acute lower extremity MSK injury, stratified by time-to-stability tertiles, was also summarized using Kaplan-Meier methods. Cox proportional hazard models were constructed to determine if longer median times to stability were associated with faster times to injury in collegiate athletes. Similar models were performed for each secondary outcome of the I-mP&R test (step latency and step length). All models were adjusted for previous musculoskeletal injury in the past two years, history of concussion, age, BMI, and sport type (contact or non-contact).

## RESULTS

Participant characteristics are listed in Table 1. At baseline, 34.0% of the sample had a history of concussion, defined as at least one lifetime concussion, and 45.0% of the sample had a previous musculoskeletal injury in the past two years (**Table 1**). Prospectively, forty-five athletes (24%; 29 F) suffered a MSK injury over an average follow-up of 198 days. Time loss from these MSK injuries ranged from three to 497 days (median (IQR) = 25.5 (390) days). Descriptive frequencies of lower extremity MSK injuries by body part are given in **Supplementary Table 2**. Ankle (29.5%), foot (13.6%), groin/hip (13.4%), and lumbar spine (11.1%) injuries were most common.

Each 250 ms increase in dual-task median time-to-stability was associated with a 37% increased risk of acute, lower-extremity MSK after adjusting for previous MSK injury in the past two years, history of concussion, age, BMI, and contact/non-contact sport (hazard ratio (HR) = 1.37, 95% confidence interval (CI) = 1.05-1.79, *p* = 0.0225; **Figure 1 right**, Table 2). Single- and dual-task step length and step latency, and single-task time-to-stability (**Figure 1, left**) were not significant predictors of acute lower extremity MSK injury risk (**Table 2**). Concussion history (*p* = 0.529), musculoskeletal injury history (*p* = 0.718), and sport type (*p* = 0.174) were not significantly different between tertiles of dual task time-to-stability (**Table 1, Supplementary Figure 2**). However, there was a significant difference in the distribution of males and females between tertiles of dual task time-to-stability (**Table 1**); a majority of females were in the slowest tertile (67.2%, 1.22-1.91 seconds) versus a majority of the males were in the fastest tertile (61.9%, 0.63-0.97 seconds).

**Figure 1.**
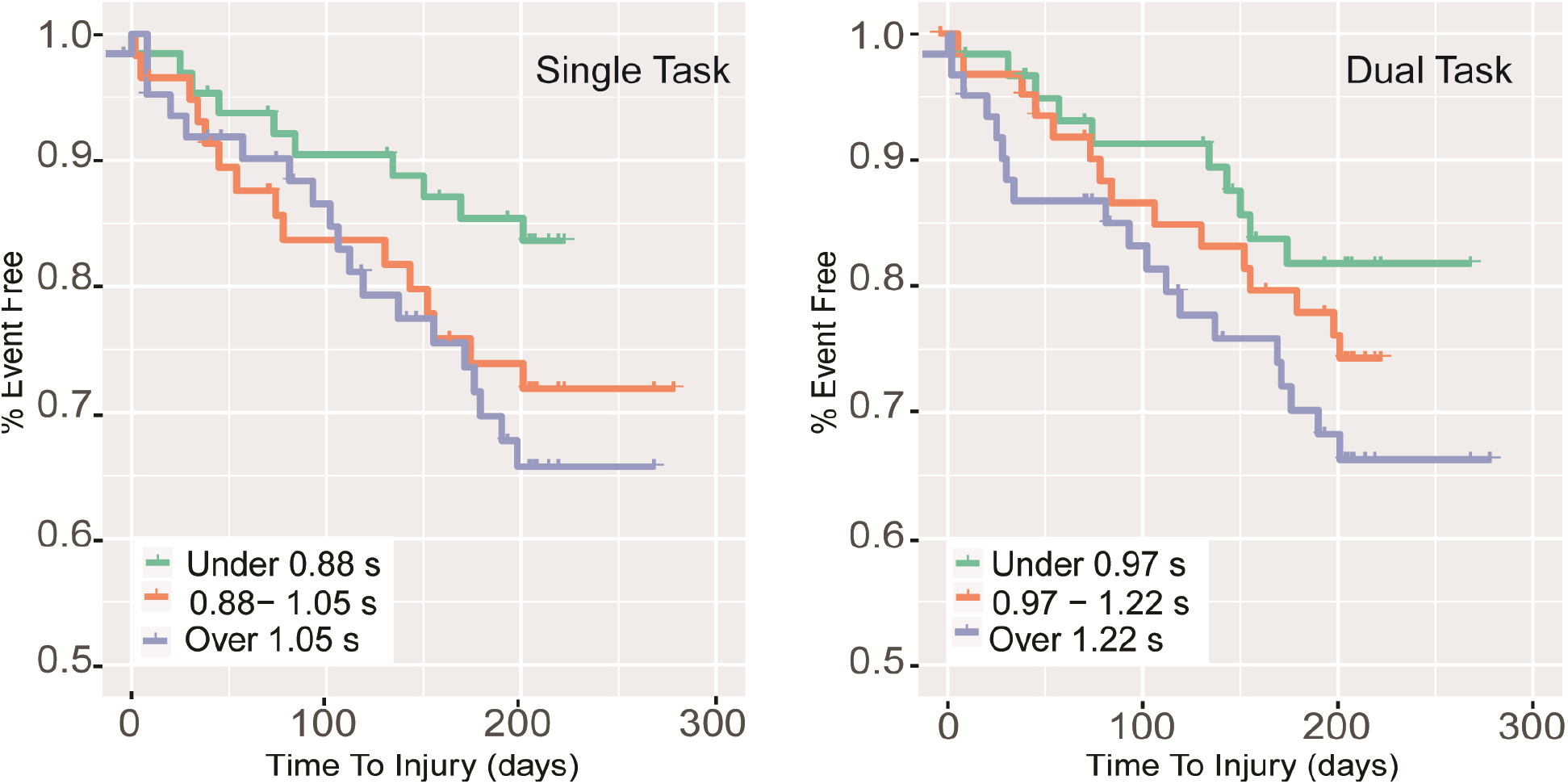
(Left) Kaplan Meier curve for time to injury from first team activity by tertile of single task median time-to-stability. (Right) Kaplan Meier curve for time to injury from first team activity by tertile of dual task median time-to-stability

**Table 2.** Adjusted hazard ratios (HR) and confidence intervals (CI)

| *Variable* |  | *HR (95% CI)* | *p value* |
| --- | --- | --- | --- |
| *Reactive Postural Responses* | *Task* |  |  |
| Time-to-stability, per 250 ms | ST | 1.29 (0.94-1.78) | 0.1205 |
| Time-to-stability, per 250 ms | DT | 1.37 (1.05-1.79) | 0.0225 |
| Step Length, per 25 cm | ST | 1.01 (1.00-1.02) | 0.2316 |
| Step Length, per 25 cm | DT | 1.01 (1.00-1.02) | 0.1595 |
| Step Latency, per 50 ms | ST | 1.12 (0.98-1.27) | 0.0856 |
| Step Latency, per 50 ms | DT | 0.92 (0.82-1.03) | 0.1667 |

Models were adjusted for previous MSK injury in the past two years, history of concussion, age, BMI, and contact/non-contact sport. The follow-up period spanned the first official team activity to March 14, 2020, when athletic activities were halted due to COVID-19. All hazard represent a one unit change unless otherwise specified. *ST*, Single Task, *DT* Dual Task, *ms* milliseconds, *cm* centimeters

## DISCUSSION

This study examined objective measures of reactive balance as predictors of risk for future acute lower extremity musculoskeletal injury. In agreement with our hypothesis we found that collegiate athletes who took longer to regain stability under dual-task conditions had greater risk for future acute MSK injury. Our results suggest that assessments that incorporate multisensory stimuli and attentional demands with precise motor responses to perform a functional movement are better indicators of a MSK injury risk, potentially because such tasks reflect the neuromechanical demands of sport.

Reactive postural responses mimic unanticipated cutting and change of direction tasks that are common in dynamic sporting environments and associated with non-contact MSK injuries. Both unanticipated tasks and reactive postural responses involve quick, time constrained responses to external stimuli that cannot be anticipated.^24^ Unanticipated change-of-direction tasks result in changes to biomechanical variables (e.g. knee abduction angle, knee flexion angle, hip and knee coupling angle) that are associated with increased anterior cruciate ligament (ACL) loads^25^ and increased ankle joint anterior shear force.^26^ Further, non-contact ACL injuries are likely to occur ∼40 ms after initial contact with the ground, when knee valgus angle increases and peak ground reaction force is 3.2 times body weight.^27^ This time frame (40 ms) is too quick to facilitate feedback driven control following an inaccurate movement - the ability to use feedforward control, encompassing the pre-perturbation neuromotor state and prediction of possibilities for future movement,^28^ to control the kinematic state of the body is therefore critical to minimizing the risk of MSK injury. Time-to-stability reflects this feedforward control; the more precise the initial foot placement (based on the kinematic state during the fall), the less time it takes to regain stability. As support is released, sensory systems (proprioceptive, visual, and vestibular) contribute to an individual’s state estimation (i.e. internal estimation of orientation and velocity).^29^ The state estimation is then used to form a feedforward, predictive model containing corrective actions (i.e. foot placement) to maintain balance. If the predictive model is incorrect, the resulting foot placement is incorrect, the individual continues to move, and the resulting TTS increases. Therefore, our results suggest that the longer TTS in athletes likely reflects an impairment in rapidly incorporate sensory information into a feedforward model for precise stepping, and whole-body, control.

The association between MSK injury and dual-task TTS, but not single-task TTS, suggests that the ability to make precise feedforward movements with simultaneous attentional demands, a common occurrence in competitive athletics,^13, 14^ is particularly important for an athlete’s health. Attentional resources are needed 1) to identify sensory cues associated with a perturbation and 2) to acquire visuospatial information about the environment to update the pre-perturbation central-set for accurate foot placement, including pre-planning for subsequent movements.^28^ While there are multiple potential mechanisms that explain dual-task interference (e.g. capacity sharing, bottleneck theory, cross-talk model),^30^ individuals who, when attentional demands are added, struggle to rapidly integrate sensory information to estimate one’s kinematic state and process the motor plan based on state estimation (the central nervous system’s estimate of the state of the motor system), are likely to make mistakes in their action (e.g., imprecise foot placement). These results are in line with previous studies examining the link between cognition and drop-jump landing biomechanics. Athletes with lower baseline neurocognitive performance exhibited greater peak vertical ground reaction force, peak anterior shear force, knee abduction moment and knee abduction angle, patterns commonly associated with ACL injury.^15^ Similarly, when cognitive load was increased during a drop-jump landing in athletes, athletes displayed greater peak knee abduction angle and lower peak knee flexion angle.^5^ The simultaneous nature of the dual motor-cognitive task appears important for assessing injury risk.

Given the abrupt conclusion of prospective injury tracking due to COVID-19, there are several limitations that are important to consider when interpreting the results of this study. First, there was a significant difference in the distribution of males and females between tertiles of dual task time-to-stability; a majority of females were in the slowest tertile, while a majority of the males were in the fastest tertile. Time-to-injury was not significantly different between sexes, but a greater proportion of females (30%) compared to males (16%) experienced an acute lower extremity MSK injury over the course of the study. We were not powered to detect sex differences and therefore cannot speculate on the meaning of this difference. However, future studies should examine such sex-based differences. Second, our protocol had a shorter follow-up time than planned because of COVID-19 (∼6 months vs. planned 12 months).

While we could have continued to follow injuries after COVID-19 forced the cancellation of all university athletic activities, such an approach would have dramatically changed the exposure rates between the time before March 14, 2020 and afterwards. We opted to maintain a shorter follow-up time, thus increasing the amount of right-censored data and limiting power. However, the fact that we still found reactive postural responses to be a significant predictor of future injury risk indicates that reactive postural responses should be considered in assessing future risk of MSK injury. Future studies should seek to confirm these results in a larger cohort with longer follow-up.

## CONCLUSION

This study demonstrates the applicability of a simple, objective measure to identify athletes at risk for prospective lower extremity MSK injury. Results from this study provide evidence to support the idea that the ability to rapidly integrate sensory information and execute a precise response, particularly under cognitive/attentional loads, is critical to minimizing injury risk.^13^ Additionally, these results show the clinical utility of longer stabilization times during the dual-task I-mP&R. Future research should focus on populations at risk of MSK injury (e.g. concussed athletes).

## Data Availability

Data produced in the present study are available upon reasonable request to the authors

## PRACTICAL IMPLICATIONS

- The instrumented, modified push-and-release predicts acute lower extremity MSK injuries in collegiate athletes.
- Time-to-stability is an objective measure of the participant’s ability to maintain or return to equilibrium after an external disturbance.
- Longer time-to-stability in reactive balance testing may reflect neuromuscular control abnormalities.
- Reactive balance testing may be able to identify athletes at risk of future musculoskeletal injury.

## Ethics Approval

Approval was granted by the Institutional Review Board of the University of Utah (Date: 2/28/2019/No: IRB_00116431)

