## Supplemental Table 1 for "Reactive Postural Responses Predict Risk For Acute Musculoskeletal Injury In Collegiate Athletes"

### SUPPLEMENTARY MATERIAL

**Supplementary Table 1.** Numbers of athletes per sport by sex.

| Sport | Female (N = 104) | Male (N = 83) |
|---|---|---|
| *Basketball* | 9 | 13 |
| *Baseball* | N/A | 12 |
| *Lacrosse* | N/A | 27 |
| *Tennis* | 3 | 5 |
| *Golf* | N/A | 7 |
| *Swimming* | 9 | 14 |
| *Skiing* | 6 | 9 |
| *Soccer* | 19 | N/A |
| *Gymnastics* | 11 | N/A |
| *Softball* | 17 | N/A |
| *Cross Country* | 4 | N/A |
| *Track* | 4 | N/A |
| *Volleyball (Indoor)* | 15 | N/A |
| *Volleyball (Beach)* | 6 | N/A |
| *Multiple* | 1 | 0 |

N = number

**Supplementary Table 2.** Injury characteristics of men and women stratified by dual-task time-to-stability tertiles.

| | *Sex* | | | | | | ***Total Injuries by Body Part*** |
|---|---|---|---|---|---|---|---|
| | ***Female*** | | | ***Male*** | | | |
| | *Tertile 1 (0.64 -1.04 sec)* | *Tertile 2 (1.04 -1.29 sec)* | *Tertile 3 (1.30-3.48 sec)* | *Tertile 1 (0.64 -1.04 sec)* | *Tertile 2 (1.04 -1.29 sec)* | *Tertile 3 (1.30-3.48 sec)* | |
| | *N = 6* | *N = 11* | *N = 12* | *N=4* | *N=5* | *N=7* | *N = 45* |
| *Body part* | | | | | | | |
| *Ankle* | 2 (33.3%) | 3 (27.3%) | 3 (25.0%) | 1 (25.0%) | 1 (20.0%) | 3 (42.9%) | 13 |
| *Ankle/Heel* | . | 1 (9.1%) | 1 (8.3%) | . | . | . | 2 |
| *Buttock* | . | . | . | . | . | 1 (14.2%) | 1 |
| *Foot* | 1 (16.7%) | 3 (27.3%) | . | 1 (25.0%) | . | 1 (14.2%) | 6 |
| *Groin/Hip* | 1 (16.7%) | 1 (9.1%) | 1 (8.3%) | 2 (50.0%) | 1 (20.0%) | . | 6 |
| *Hamstring* | . | 1 (9.1%) | . | . | . | . | 1 |
| *Knee* | . | . | 4 (33.3%) | . | . | . | 4 |
| *Lower leg* | 1 (16.7%) | . | . | . | . | . | 1 |
| *Quadriceps* | . | 1 (9.1%) | 1 (8.3%) | . | 1 (20.0%) | 1 (14.2%) | 4 |
| *Thigh* | 1 (16.7%) | . | . | . | . | . | 1 |
| *Lumbar Spine* | . | 1 (9.1%) | 2 (16.7%) | . | 1 (20.0%) | 1 (14.2%) | 5 |
| *Unknown* | | | | | 1 (20.0%) | | 1 |
| ***% by Sex in each tertile*** | 20.7% | 37.9% | 41.3% | 25.0% | 31.2% | 43.8% | |

Percents represent the proportion of individuals among a given tertile with an injury at a given body part or sex.
*Sec* seconds, *N* = number

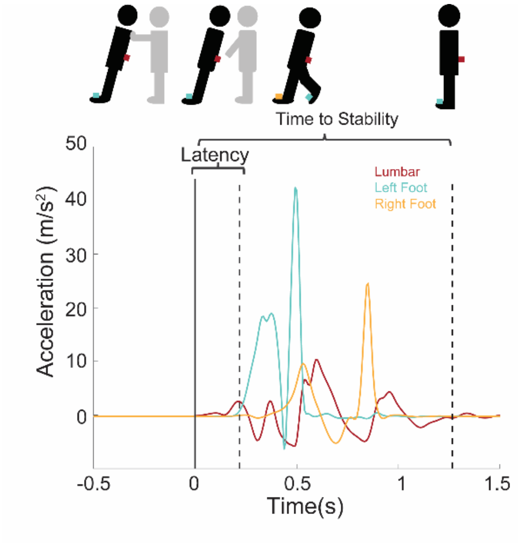


**Supplementary Figure 1.** Representative acceleration profile for the lumbar (red), left foot (green), and right foot (yellow) sensors. Stick figures represent time course of push and release; Grey stick figure represents administrator. Black stick figure represents participant. Colored boxes represent sensor placement; red = lumbar, green = left foot, yellow = right foot.

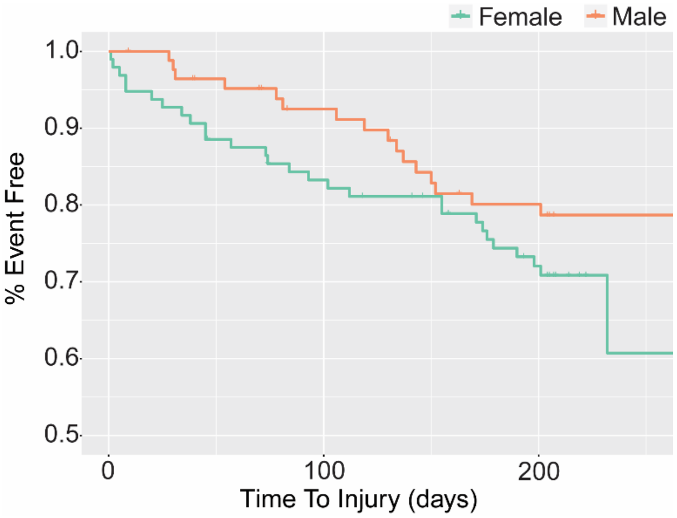


**Supplementary Figure 2.** Kaplan Meier curve for time to injury from first team activity by sex. Kaplan Meier estimates did not significantly differ by sex (Log Rank Test, $p$=0.19)
